# Investigating the intention to receive the COVID-19 vaccination in Macao: implications for vaccination strategies

**DOI:** 10.1101/2021.06.11.21258734

**Authors:** Carolina Oi Lam Ung, Yuanjia Hu, Hao Hu, Ying Bian

## Abstract

**Importance:** Understanding the intention of receiving COVID-19 vaccines is important for informing effective vaccination strategies especially for areas with low incidence.

**Objective:** This study aimed to investigate the intention to receive COVID-19 vaccination, identify the key influencing factors, and determine the most important intention predictors using a theoretically principled model.

**Design, setting, and participants:** This was a cross-sectional online survey study hosted by Survey Monkey and implemented for 10 days from May 14 2021. People who aged 18 years or above and had been residing in Macao for 12 months prior to the study were recruited through social media.

**Main outcomes and measures:** Intention, the constructs of protection motivation theory (perceived severity, perceived susceptibility, maladaptive response reward, self-efficacy, response-efficacy, and response cost), constructs of health belief model (cues to action), social attitude, social norm, past experience and information seeking behavior, in addition to demographic variables.

**Results:** Of the 552 respondents, 79.5% aged between 25 and 54 years old, 59.4% were female, and 88% had a bachelor degree or above. Overall, 62.3% of the respondents indicated their intention while 19.2% were hesitant and 18.5% did not have any intention. While 67.0% believed COVID-19 infection was life-threatening, only 19.0% thought they were at risk. Control variables such as age, gender, education level, and having travel plans were significantly correlated with intention. Significant associations were found between intention and all the measures (p<0.05). The most important positive predictors of intention were “*being able to make arrangement to receive the vaccine*” (β = 0.333, P <0.001), “*a sense of social responsibility*” (β = 0.326, P < 0.001), and “*time off from work after vaccination*” (β = 0.169, P <0.001), whereas “*concerns over vaccine safety*” (β = - 0.124, P < 0.001) and “*relying on online resources for vaccine information*” (β = -0.065, P <0.05) were negative predictors. Perceived severity was not a predictor of intention.

**Conclusion and relevance:** Multi-component strategies that address various factors affecting intention are needed to formulate effective interventions. Health literacy, vaccination convenience, social responsibility, reasonable incentives and well-informed risk and benefit analysis are recommended consideration for future vaccination campaigns.

**Key Points:** *Question:* What is the intention to receive COVID-19 vaccination in Macao and the influencing factors?

*Findings:* A cross-sectional study involving 552 respondents found that the intention rate of COVID-19 vaccination was 62.3% in Macao. Positive predictors of intention included being able to make arrangement to receive the vaccine, a sense of social responsibility, and an offer of time off from work after vaccination. Concerns over vaccine safety and relying on online resources for vaccine information were negative predictors.

*Meaning:* Intention to receive COVID-19 vaccination is multifactorial requiring multi-component strategies to promote vaccine uptake.

## Introduction

The impact of COVID-19 pandemic on human health, economy and societal activities is unprecedented. Health systems continue to endure massive challenges in preventing and managing the infection with COVID-19 and its variants, and be stretched to deliver effective public health measures.^1^

One of the critical public health interventions to mitigate COVID-19 pandemic at the population level is for individuals to get vaccinated with COVID-19 vaccines. Since the release of the genetic sequence of SARS-CoV-2 in January 2020, the R&D activity to develop a vaccine against the disease has been intense across the globe.^2^ As of June 1 2021, at least 102 vaccines were in clinical development.^3^ Evidence about the safety and efficacy of COVID-19 vaccines in preventing symptomatic and asymptomatic COVID-19 infections, related hospitalizations, severe disease, death, and protection against variant strains is acumulating.^4-6^ Drug regulatory authorities and the World Health Organization have successively listed a number of new COVID-19 vaccine candidates for emergency use, and many governments rushed to distribute the vaccines to their people. ^7-10^

The speedy development, approval and distribution of COVID-19 vaccines, however, do not necessarily translate into full-range vaccine uptake. As of June 3 2021, only 41.2% of the total population in the US were fully vaccinated^11^ ; in China, 723.5 million doses were administered on the same day^12^ but to vaccinate 40% of population by end-July was a speculation^13^. Vaccination rates also varied largely among other developed countries (40.1% in the UK, 20.1% in Germany, 17.6% in France, 6.4% in Canada, 3.14% in Japan).^14^ Besides availability, accessibility and affordability, vaccine hesitancy affecting an individual’s decision-making process about getting vaccinated is a critical challenge to national vaccination programs.^15,16^ Vaccine hesitancy refers to “*delay in acceptance or refusal of vaccination despite availability of vaccination services*”, and a high rate of hesitancy or a low rate of intention would undermine the level of demand for a vaccine.^15^ Considering the far-reaching goal of 60-75% vaccination rate to achieve herd immunity^17,18^, it is paramount to have a good understanding of the vaccination intention in order to encourage vaccine uptake.

The decision about taking a COVID-19 vaccine is multifactorial and context-specific varying across people, time, place and vaccines. ^15^ There is an ongoing need to investigate the factors contributing to vaccine hesitancy or affecting the intention to get vaccinated against COVID-19. This is particularly important for areas whereby intention may be hampered by low incidence of COVID-19 cases. Therefore, the aim of this study was to investigate the intention to get vaccinated against COVID-19 and the associations between such intentions and theoretically principled and sociodemographic factors in Macao.

## Methods

An online, cross-sectional survey method was applied in this study. The project has been approved by the Panel on Research Ethics of the University of Macau in May 2021 (SSHRE21-APP018-ICMS). As indicated in the Participant Information Statement, it was assumed that, by completing and submitting the survey online, participants agreed to take part in the research study. This study followed the American Association for Public Opinion Research (AAPOR) reporting guideline.

Macau is a city with a population of around 690,000 over 32.9km^2^ ranked one of the most densely populated places in the world. As of June 6 2021, there were only 51 imported cases in the city all of whom fully recovered, and no signs of community transmissions.^19^ The percentage of population fully vaccinated was around 10%. The target population in this study was people aged 18 years or above who had resided in Macao in the past 12 months. A minimum sample size of 384 respondents would provide a target 5% margin of error for population percentage estimates with a level of 95% confidence.

The online questionnaire, hosted by Survey Monkey, was open on from May 14 2021 and closed on May 23 2021 when no new responses were received for 24 hours. Invitations to participate in the study were distributed through social media like Facebook and WeChat, and news media social platform. Given the popularity of the social media among the Macao population and the need to maintain social distancing, social media and online communication platform was chosen. Simplified-snowball sampling technique was also used to recruit more people from respondents’ contact. The survey took less than 8 min to complete on average.

### Measures

The design of the questionnaire was informed by (1) the current literature on intention of COVID-19 vaccination^20-22^, (2) the health belief model^23^ and the protection motivation theory^24^, and (3) a clinician involved in the local COVID-19 vaccination campaign. The schematic diagram of the theoretically principled constructs applied to the intention to receive COVID-19 vaccination in this study is provided in Figure 1.

**Figure 1.**
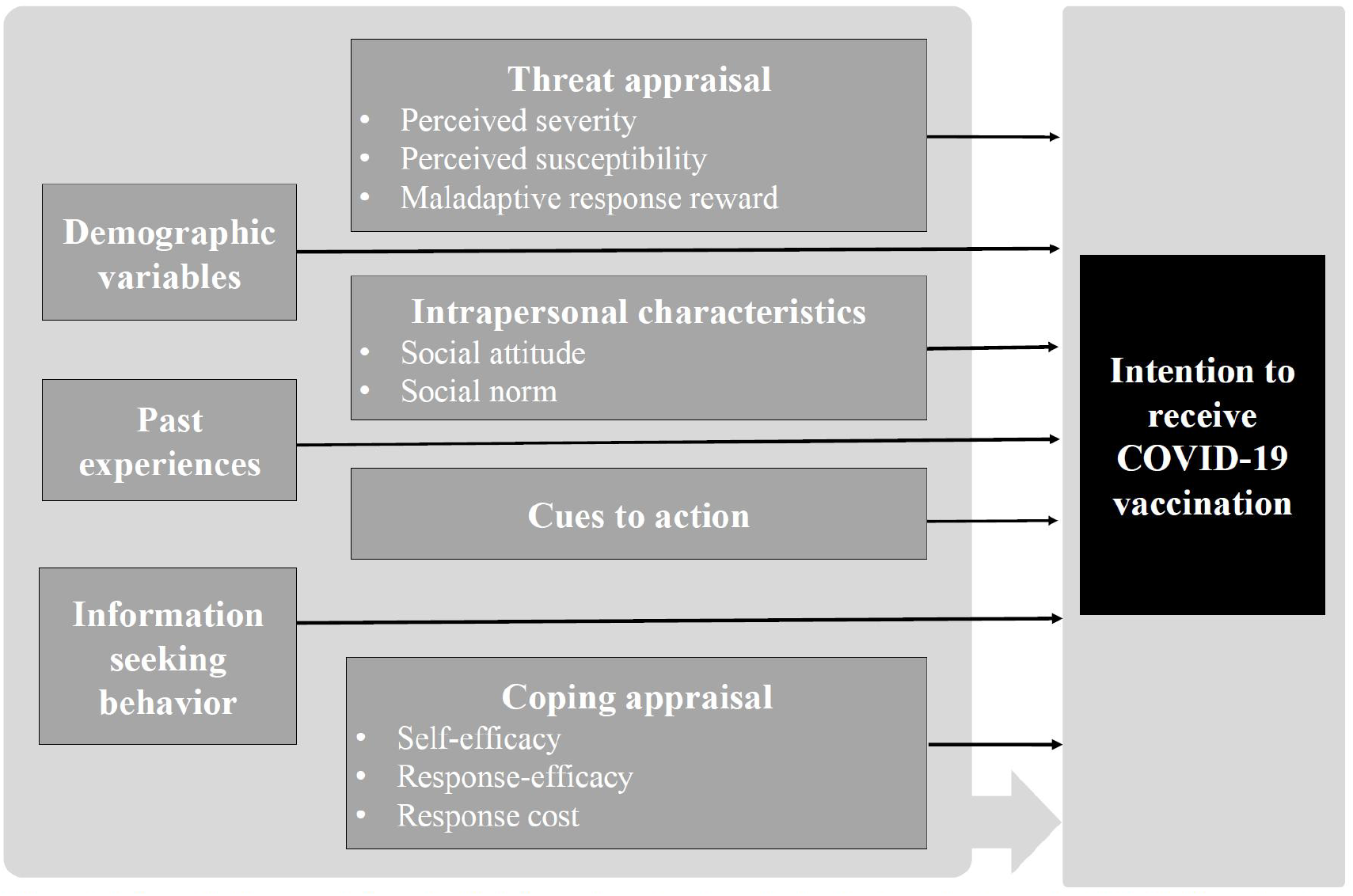
Schematic diagram of theoretically-informed constructs applied to the intention to receive COVID-19 vaccination.

(Figure 1. Schematic diagram of theoretically-principled constructs applied to the intention to receive COVID-19 vaccination)

The questionnaire mainly comprised of 2 sections: Section A collect respondents’ demographic information and Section B asked respondents to rate their level of agreement using a 5-point Likert scale (1=strongly disagree, 2=disagree, 3=not sure, 4=agree, and 5=strongly agree) their intention and 28 items representing 11 constructs: perceived severity, perceived susceptibility, maladaptive response rewards, self-efficacy, response-efficacy, response costs, social attitude, social norms, past experiences, information seeking behavior, and cues for action. At the conclusion of the study, participants were invited to provide additional feedback via a free-text response box.

The questionnaire was pilot-tested twice (2 doctors, 2 pharmacists and 2 researchers in Round 1, and 10 individuals in Round 2) following the concepts described by Leavy^25^ to assess content validity, content consistency, and comprehension, defective questions and the time needed to complete.

### Statistical analysis

The data was analyzed using the Statistical Package for Social Sciences (SPSS) version 24 software for Windows. In addition to descriptive statistics (frequencies, means, and standard deviations), Pearson chi-square test was used to compare the differences in COVID-19 vaccination intentions among subgroups, Spearman’s rho was used to test the correlation of intention with the variables, and multiple linear regression analysis to identity predictors of intention. Whenever the P-value is found to be smaller than 0.05, the association would be considered statistically significant at a confidence level of 95%.

## Results

### Respondents’ characteristics and the differences in the intention among subgroups

A total of 552 valid responses were received. As shown in Table 1, 79.5% aged between 25 and 54 years old, 59.4% were female, 74.8% were local residents, 88% had a bachelor degree or above and 64.5% planned to travel in and out of Macao within the next 6 months. In general, the respondents were broadly representative of the population in Macao in terms of age and types of residency, but the proportion of female and people having higher levels of education were higher than intended.

**Table 1:**
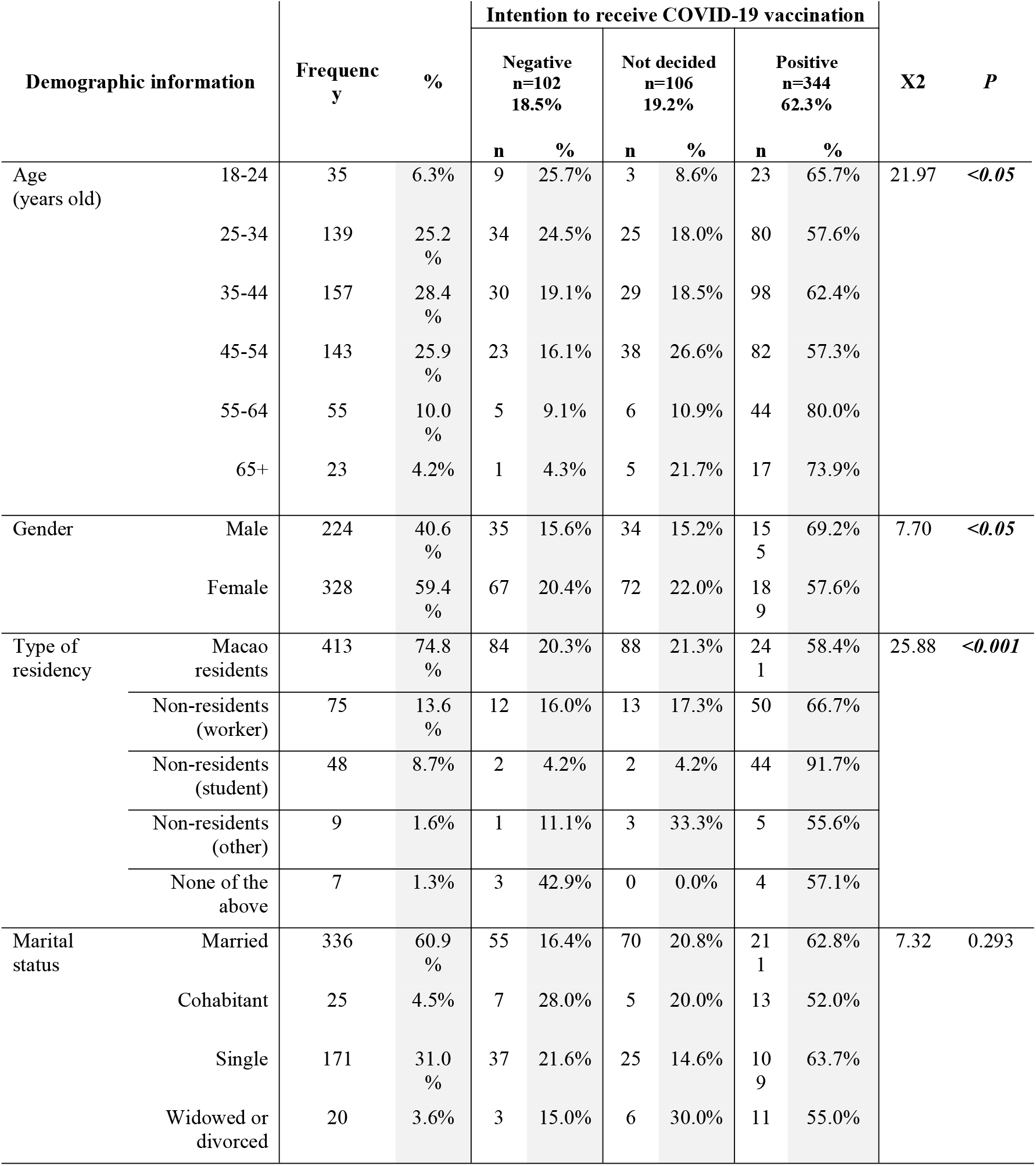
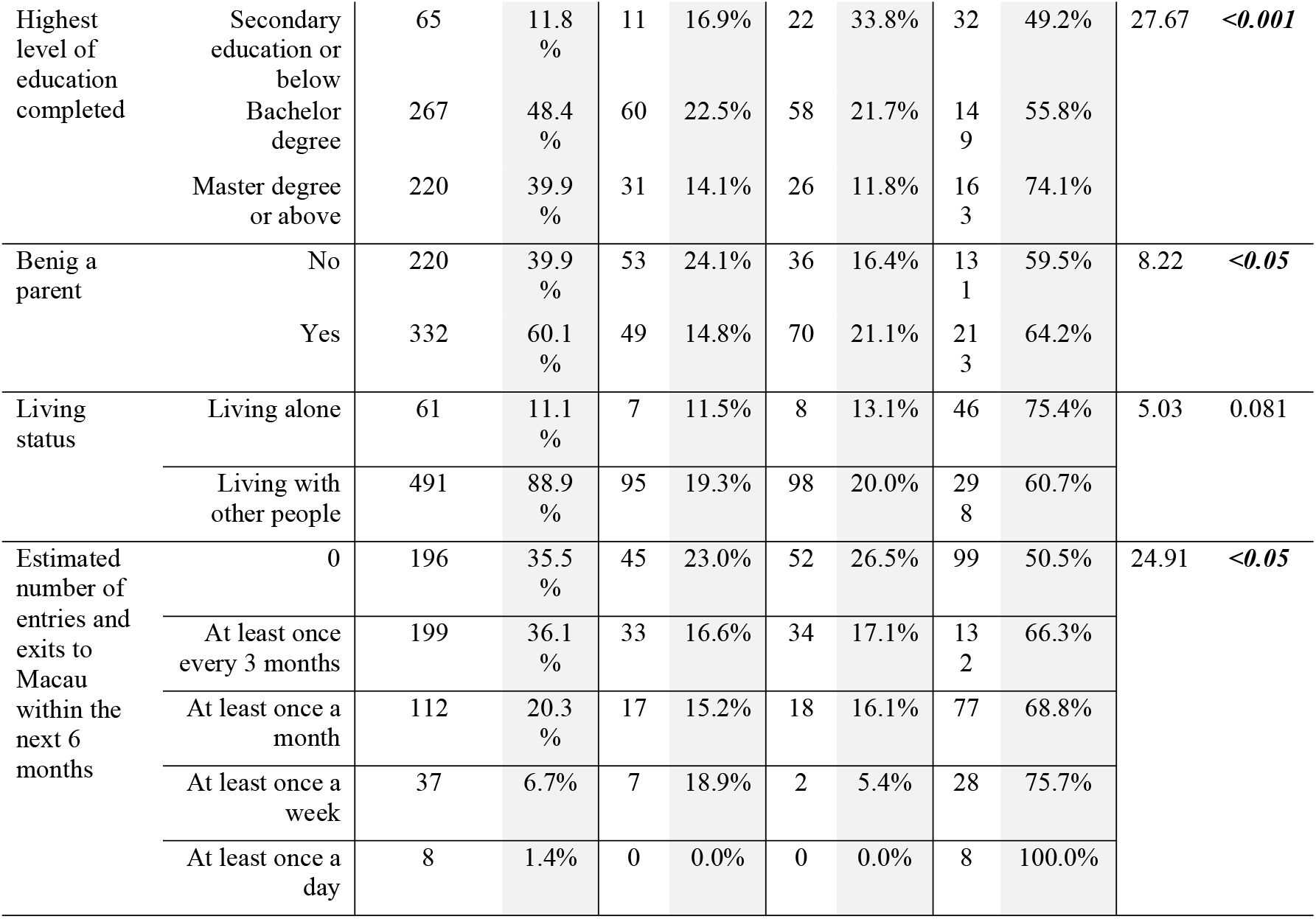
Respondents’ demographic information (n=552)

### Intention to receive COVID-19 vaccination

Overall, 62.3% of the respondents indicated their intention to receive COVID-19 vaccination, while 19.2% were hesitant and 18.5% did not want to get vaccinated at all. The intention rate within each of the subgroup is shown in Figure 2. According to the Pearson Chi-square test results (Table 1), statistically significant differences in the intention were observed among the subgroups of age, gender, residency types, education level, parental status and having plans to travel in and out of Macao.

**Figure 2.**
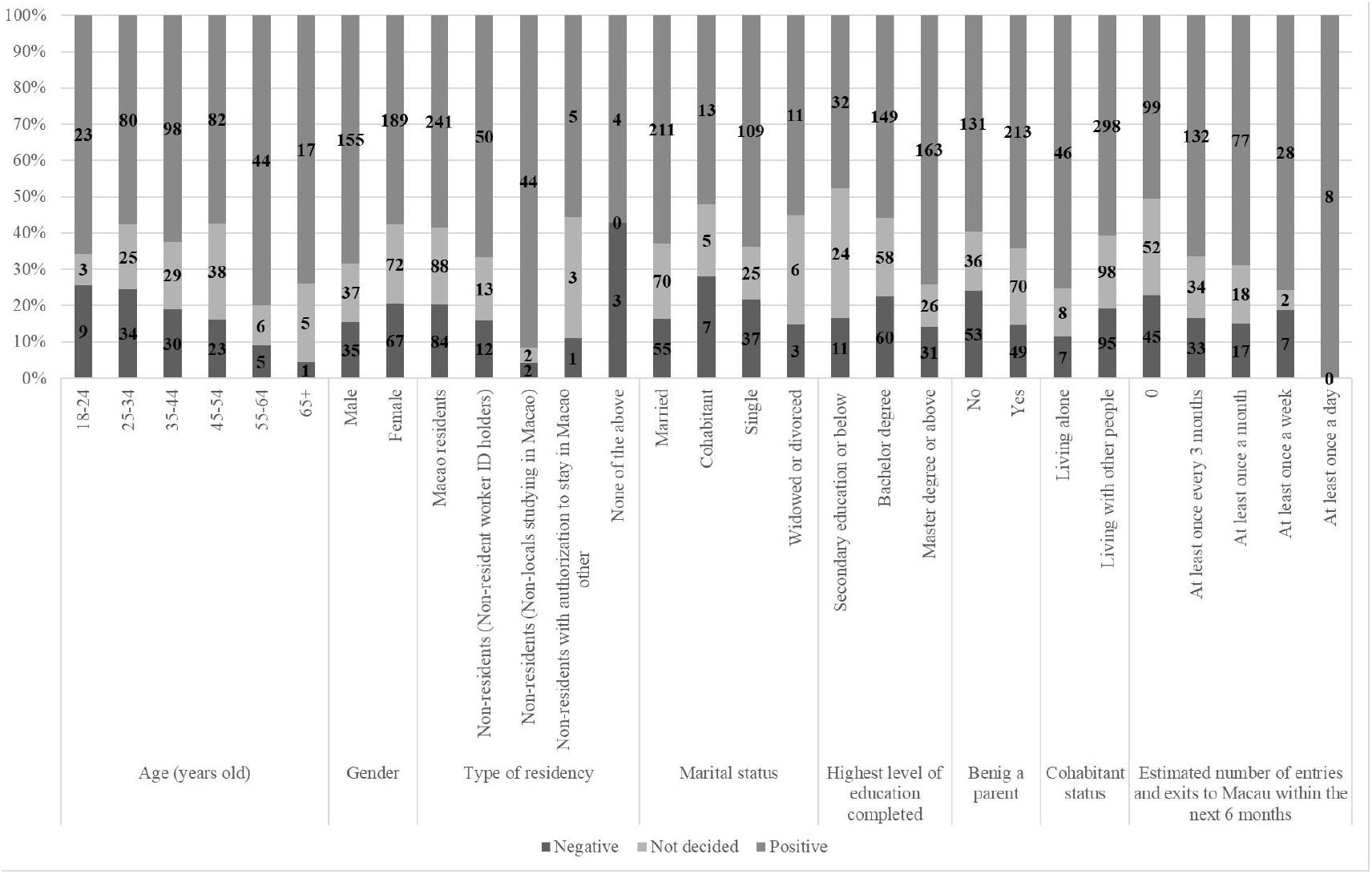
Intention of getting COVID-19 vaccination among subgroups (%)

(Figure 2. Intention of getting COVID-19 vaccination among subgroups (%))

### Respondents’ perception about the measurements

Descriptive statistics for items assessing factors related to intention are reported in Table 2. The results of Spearman’s rho suggested that almost all of the items were significantly associated with respondents’ level of intention to receive COVID-19 vaccination.

**Table 2.**
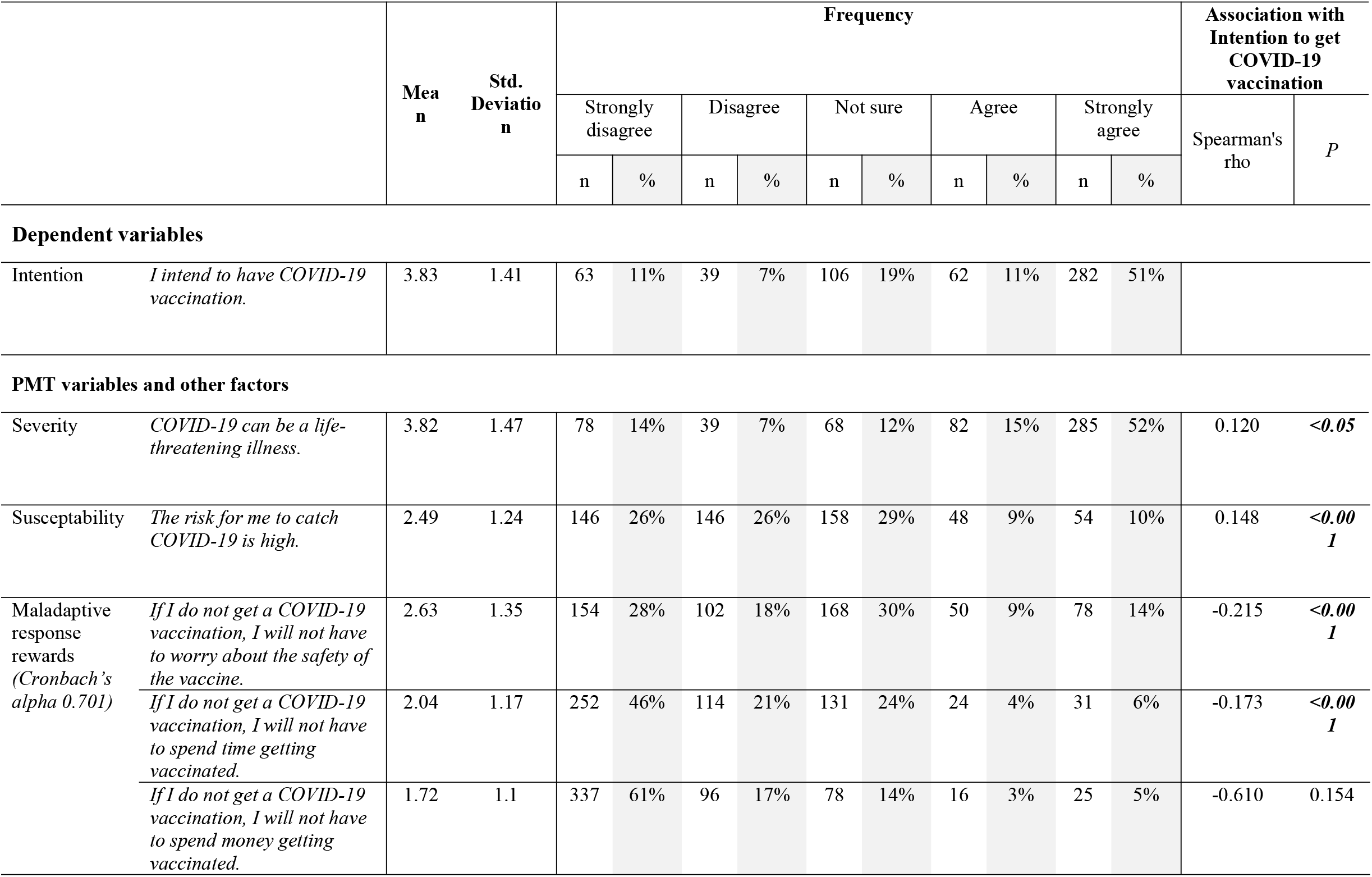
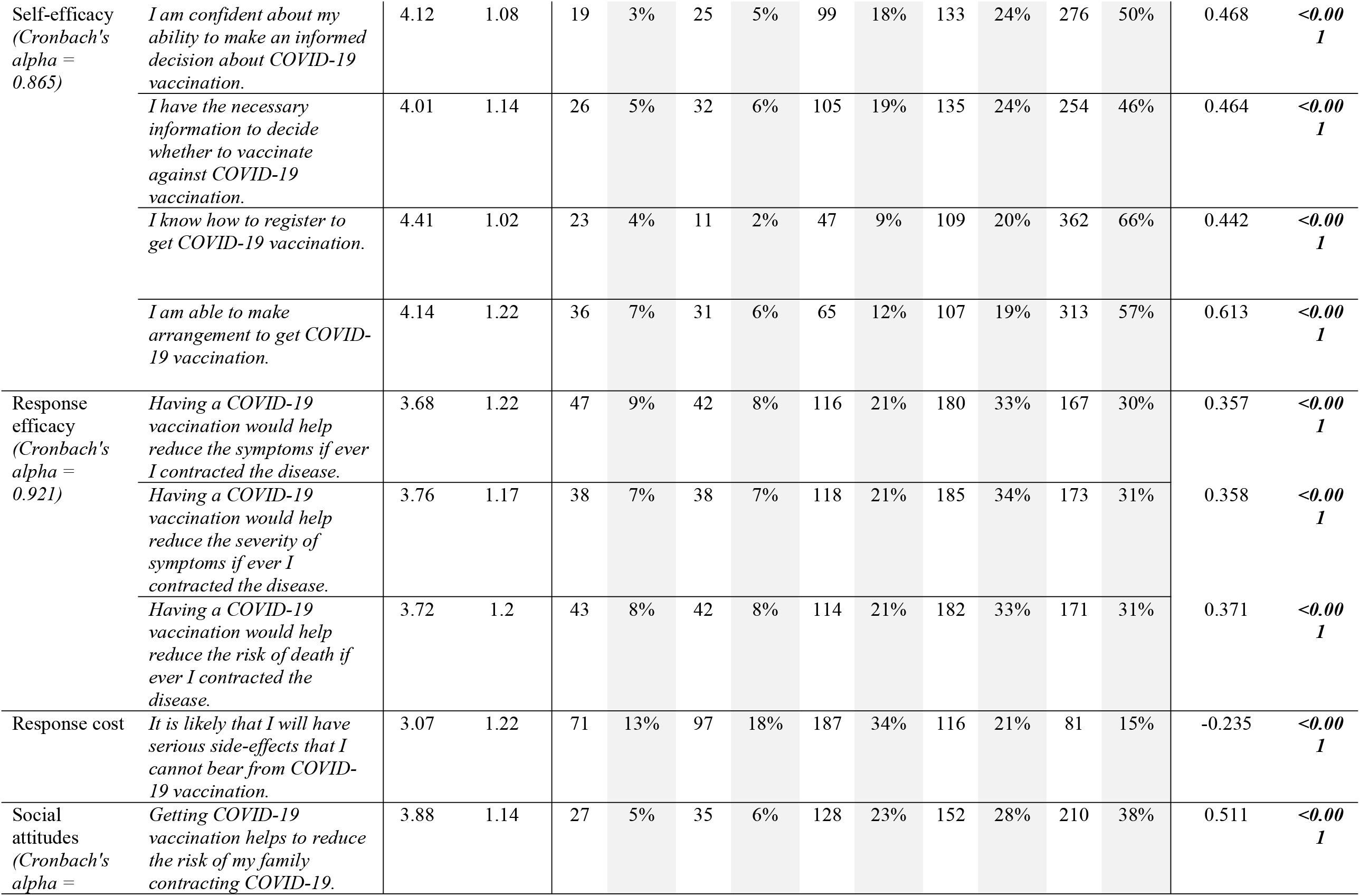
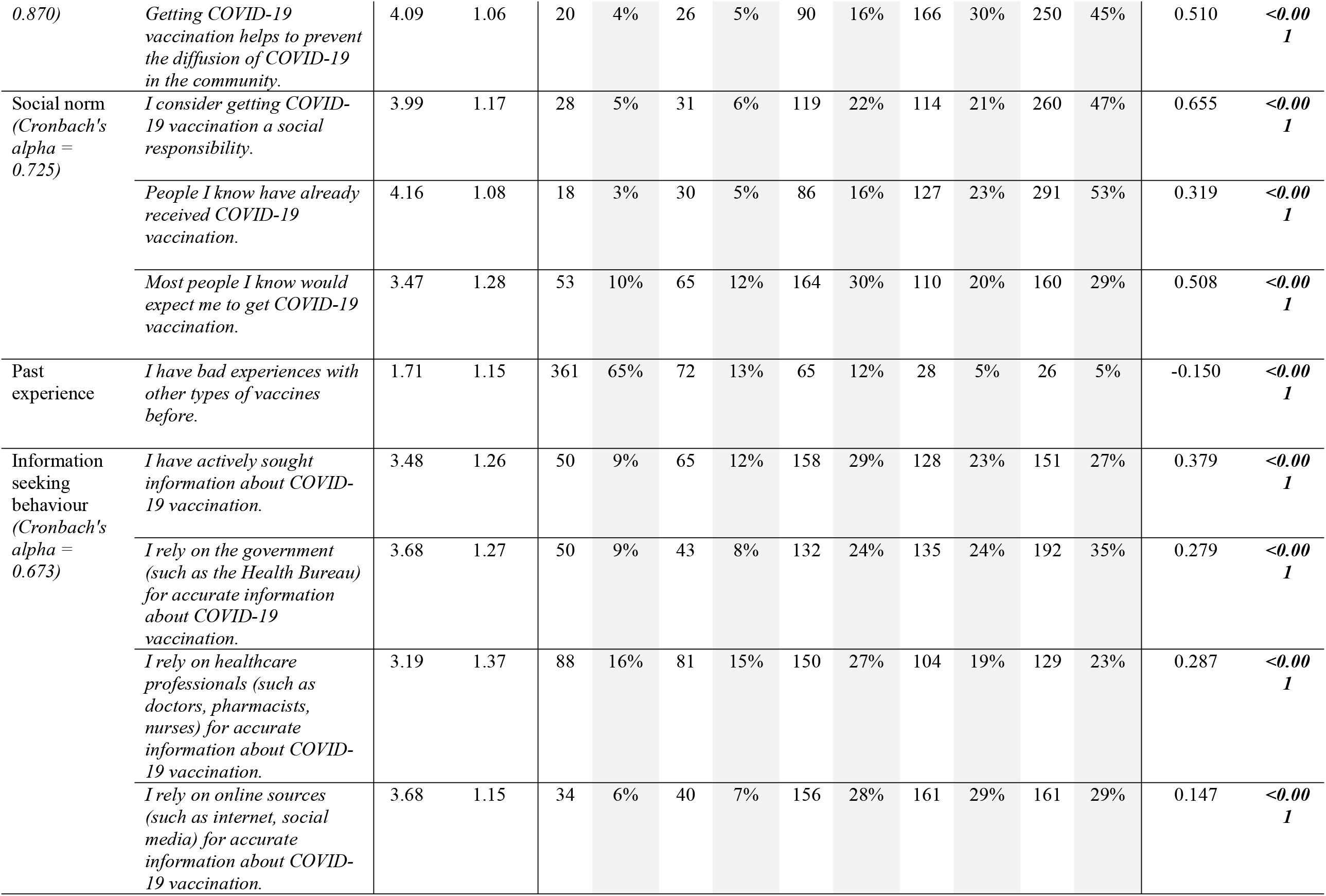
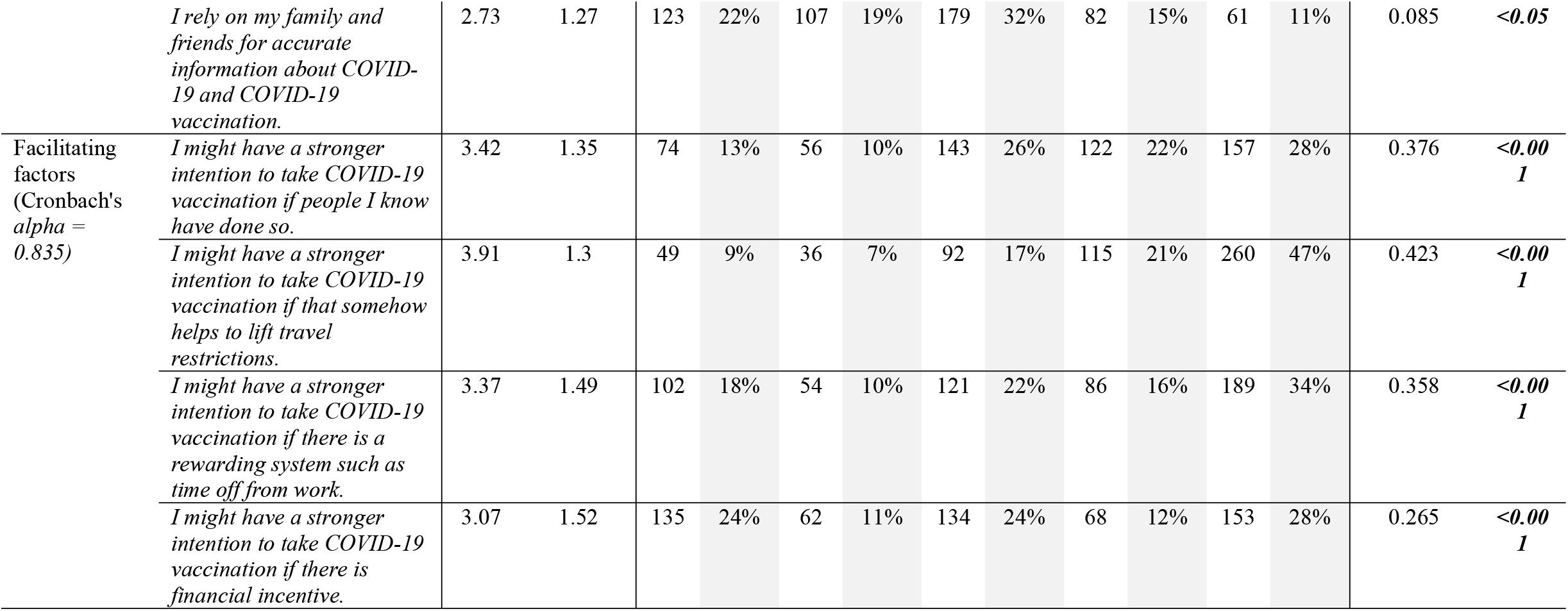
Measurement of variables and other factors.

|  |  | Mea<br>n | Std.<br>Deviation | Frequency |  |  |  |  |  |  |  |  |  | Association with<br>Intention to get<br>COVID-19<br>vaccination |  |
| --- | --- | --- | --- | --- | --- | --- | --- | --- | --- | --- | --- | --- | --- | --- | --- |
|  |  |  |  | Strongly<br>disagree |  | Disagree |  | Not sure |  | Agree |  | Strongly<br>agree |  | Spearman's<br>rho | P |
|  |  |  |  | n | % | n | % | n | % | n | % | n | % |  |  |
| Dependent variables |  |  |  |  |  |  |  |  |  |  |  |  |  |  |  |
| Intention | *I intend to have COVID-19 vaccination.* | 3.83 | 1.41 | 63 | 11% | 39 | 7% | 106 | 19% | 62 | 11% | 282 | 51% |  |  |
| PMT variables and other factors |  |  |  |  |  |  |  |  |  |  |  |  |  |  |  |
| Severity | *COVID-19 can be a life-threatening illness.* | 3.82 | 1.47 | 78 | 14% | 39 | 7% | 68 | 12% | 82 | 15% | 285 | 52% | 0.120 | **<0.05** |
| Susceptability | *The risk for me to catch COVID-19 is high.* | 2.49 | 1.24 | 146 | 26% | 146 | 26% | 158 | 29% | 48 | 9% | 54 | 10% | 0.148 | **<0.001** |
| Maladaptive response rewards<br>(Cronbach's alpha 0.701) | *If I do not get a COVID-19 vaccination, I will not have to worry about the safety of the vaccine.* | 2.63 | 1.35 | 154 | 28% | 102 | 18% | 168 | 30% | 50 | 9% | 78 | 14% | -0.215 | **<0.001** |
|  | *If I do not get a COVID-19 vaccination, I will not have to spend time getting vaccinated.* | 2.04 | 1.17 | 252 | 46% | 114 | 21% | 131 | 24% | 24 | 4% | 31 | 6% | -0.173 | **<0.001** |
|  | *If I do not get a COVID-19 vaccination, I will not have to spend money getting vaccinated.* | 1.72 | 1.1 | 337 | 61% | 96 | 17% | 78 | 14% | 16 | 3% | 25 | 5% | -0.610 | 0.154 |
| Self-efficacy<br>(Cronbach's<br>alpha =<br>0.865) | *I am confident about my ability to make an informed decision about COVID-19 vaccination.* | 4.12 | 1.08 | 19 | 3% | 25 | 5% | 99 | 18% | 133 | 24% | 276 | 50% | 0.468 | **<0.001** |
|  | *I have the necessary information to decide whether to vaccinate against COVID-19 vaccination.* | 4.01 | 1.14 | 26 | 5% | 32 | 6% | 105 | 19% | 135 | 24% | 254 | 46% | 0.464 | **<0.001** |
|  | *I know how to register to get COVID-19 vaccination.* | 4.41 | 1.02 | 23 | 4% | 11 | 2% | 47 | 9% | 109 | 20% | 362 | 66% | 0.442 | **<0.001** |
|  | *I am able to make arrangement to get COVID-19 vaccination.* | 4.14 | 1.22 | 36 | 7% | 31 | 6% | 65 | 12% | 107 | 19% | 313 | 57% | 0.613 | **<0.001** |
| Response efficacy<br>(Cronbach's<br>alpha =<br>0.921) | *Having a COVID-19 vaccination would help reduce the symptoms if ever I contracted the disease.* | 3.68 | 1.22 | 47 | 9% | 42 | 8% | 116 | 21% | 180 | 33% | 167 | 30% | 0.357 | **<0.001** |
|  | *Having a COVID-19 vaccination would help reduce the severity of symptoms if ever I contracted the disease.* | 3.76 | 1.17 | 38 | 7% | 38 | 7% | 118 | 21% | 185 | 34% | 173 | 31% | 0.358 | **<0.001** |
|  | *Having a COVID-19 vaccination would help reduce the risk of death if ever I contracted the disease.* | 3.72 | 1.2 | 43 | 8% | 42 | 8% | 114 | 21% | 182 | 33% | 171 | 31% | 0.371 | **<0.001** |
| Response cost | *It is likely that I will have serious side-effects that I cannot bear from COVID-19 vaccination.* | 3.07 | 1.22 | 71 | 13% | 97 | 18% | 187 | 34% | 116 | 21% | 81 | 15% | -0.235 | **<0.001** |
| Social attitudes<br>(Cronbach's<br>alpha = | *Getting COVID-19 vaccination helps to reduce the risk of my family contracting COVID-19.* | 3.88 | 1.14 | 27 | 5% | 35 | 6% | 128 | 23% | 152 | 28% | 210 | 38% | 0.511 | **<0.001** |
| 0.870) | *Getting COVID-19 vaccination helps to prevent the diffusion of COVID-19 in the community.* | 4.09 | 1.06 | 20 | 4% | 26 | 5% | 90 | 16% | 166 | 30% | 250 | 45% | 0.510 | **<0.001** |
| Social norm<br>(Cronbach's<br>alpha =<br>0.725) | *I consider getting COVID-19 vaccination a social responsibility.* | 3.99 | 1.17 | 28 | 5% | 31 | 6% | 119 | 22% | 114 | 21% | 260 | 47% | 0.655 | **<0.001** |
|  | *People I know have already received COVID-19 vaccination.* | 4.16 | 1.08 | 18 | 3% | 30 | 5% | 86 | 16% | 127 | 23% | 291 | 53% | 0.319 | **<0.001** |
|  | *Most people I know would expect me to get COVID-19 vaccination.* | 3.47 | 1.28 | 53 | 10% | 65 | 12% | 164 | 30% | 110 | 20% | 160 | 29% | 0.508 | **<0.001** |
| Past<br>experience | *I have bad experiences with other types of vaccines before.* | 1.71 | 1.15 | 361 | 65% | 72 | 13% | 65 | 12% | 28 | 5% | 26 | 5% | -0.150 | **<0.001** |
| Information<br>seeking<br>behaviour<br>(Cronbach's<br>alpha =<br>0.673) | *I have actively sought information about COVID-19 vaccination.* | 3.48 | 1.26 | 50 | 9% | 65 | 12% | 158 | 29% | 128 | 23% | 151 | 27% | 0.379 | **<0.001** |
|  | *I rely on the government (such as the Health Bureau) for accurate information about COVID-19 vaccination.* | 3.68 | 1.27 | 50 | 9% | 43 | 8% | 132 | 24% | 135 | 24% | 192 | 35% | 0.279 | **<0.001** |
|  | *I rely on healthcare professionals (such as doctors, pharmacists, nurses) for accurate information about COVID-19 vaccination.* | 3.19 | 1.37 | 88 | 16% | 81 | 15% | 150 | 27% | 104 | 19% | 129 | 23% | 0.287 | **<0.001** |
|  | *I rely on online sources (such as internet, social media) for accurate information about COVID-19 vaccination.* | 3.68 | 1.15 | 34 | 6% | 40 | 7% | 156 | 28% | 161 | 29% | 161 | 29% | 0.147 | **<0.001** |
|  | *I rely on my family and friends for accurate information about COVID-19 and COVID-19 vaccination.* | 2.73 | 1.27 | 123 | 22% | 107 | 19% | 179 | 32% | 82 | 15% | 61 | 11% | 0.085 | **<0.05** |
| Facilitating factors<br>(Cronbach's alpha = 0.835) | *I might have a stronger intention to take COVID-19 vaccination if people I know have done so.* | 3.42 | 1.35 | 74 | 13% | 56 | 10% | 143 | 26% | 122 | 22% | 157 | 28% | 0.376 | **<0.001** |
|  | *I might have a stronger intention to take COVID-19 vaccination if that somehow helps to lift travel restrictions.* | 3.91 | 1.3 | 49 | 9% | 36 | 7% | 92 | 17% | 115 | 21% | 260 | 47% | 0.423 | **<0.001** |
|  | *I might have a stronger intention to take COVID-19 vaccination if there is a rewarding system such as time off from work.* | 3.37 | 1.49 | 102 | 18% | 54 | 10% | 121 | 22% | 86 | 16% | 189 | 34% | 0.358 | **<0.001** |
|  | *I might have a stronger intention to take COVID-19 vaccination if there is financial incentive.* | 3.07 | 1.52 | 135 | 24% | 62 | 11% | 134 | 24% | 68 | 12% | 153 | 28% | 0.265 | **<0.001** |

While respondents rated severity highly (mean=3.82±1.47), the perceived susceptibility was much lower (mean=2.49±1.24). Regarding maladaptive responses, respondents rated the money they had to spend on COVID-19 vaccination the least concern (mean=1.72±1.10). The rating for self-efficacy was high especially about their ability to register for COVID-19 vaccination (mean=4.41±1.02) and make appropriate arrangements (mean=4.14±1.22). Their ratings on the response-efficacy was relatively low regarding the effect of reducing symptoms (mean=3.68±1.22). More than one-third of the respondents believed that it would be likely for them to experience unbearable side effects after COVID-19 vaccination. In terms of social attitudes, the respondents recognized the effectiveness of COVID-19 vaccination on the community (mean=4.09±1.06) more than that on their family (mean=3.88±1.14). The respondents thought highly of getting COVID-19 vaccination as a social responsibility (mean=3.99±1.17). In terms of the source of information about COVID-19, they relied mostly on the government (mean=3.68±1.27) and least on family and friends (mean=2.73±1.27). Regarding cues for action, the respondents rated loosening of travel restrictions as a result of COVID-19 vaccination the highest (mean=3.91±1.30) and financial incentive the lowest (mean=3.07±1.52).

### Results of multiple linear regressions

All factors found to be significantly associated with intention in Table 1 and 2 were analyzed as the independent variables, and the intention as the dependent variable. As shown in Table 3, the model is able to explain 57.9 % of the variance in intention to receive COVID-19 vaccination. In addition to age, 8 items from 7 constructs were shown to be predictors of intention, including 5 positive predictors and 3 negative predictors.

**Table 3.**
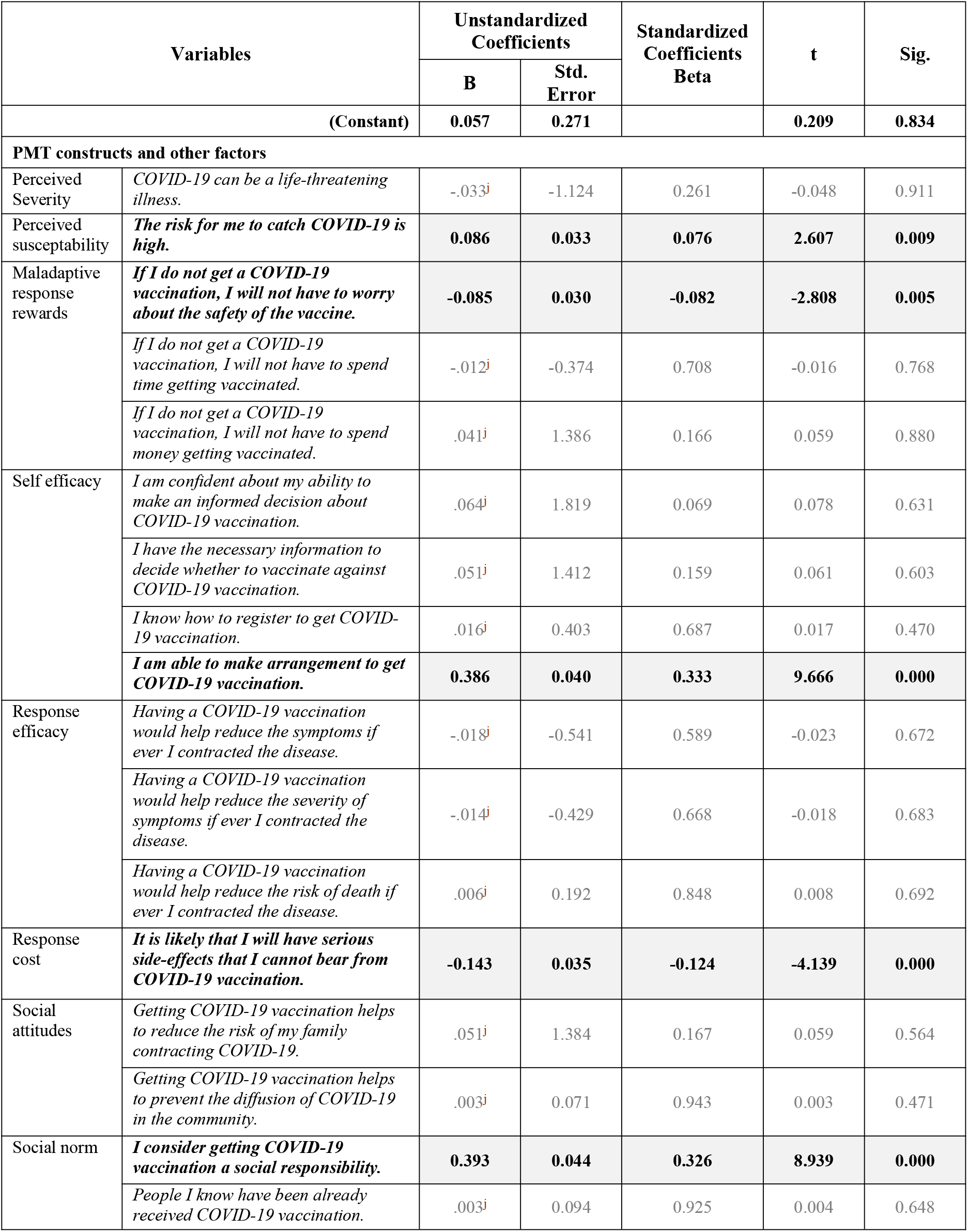
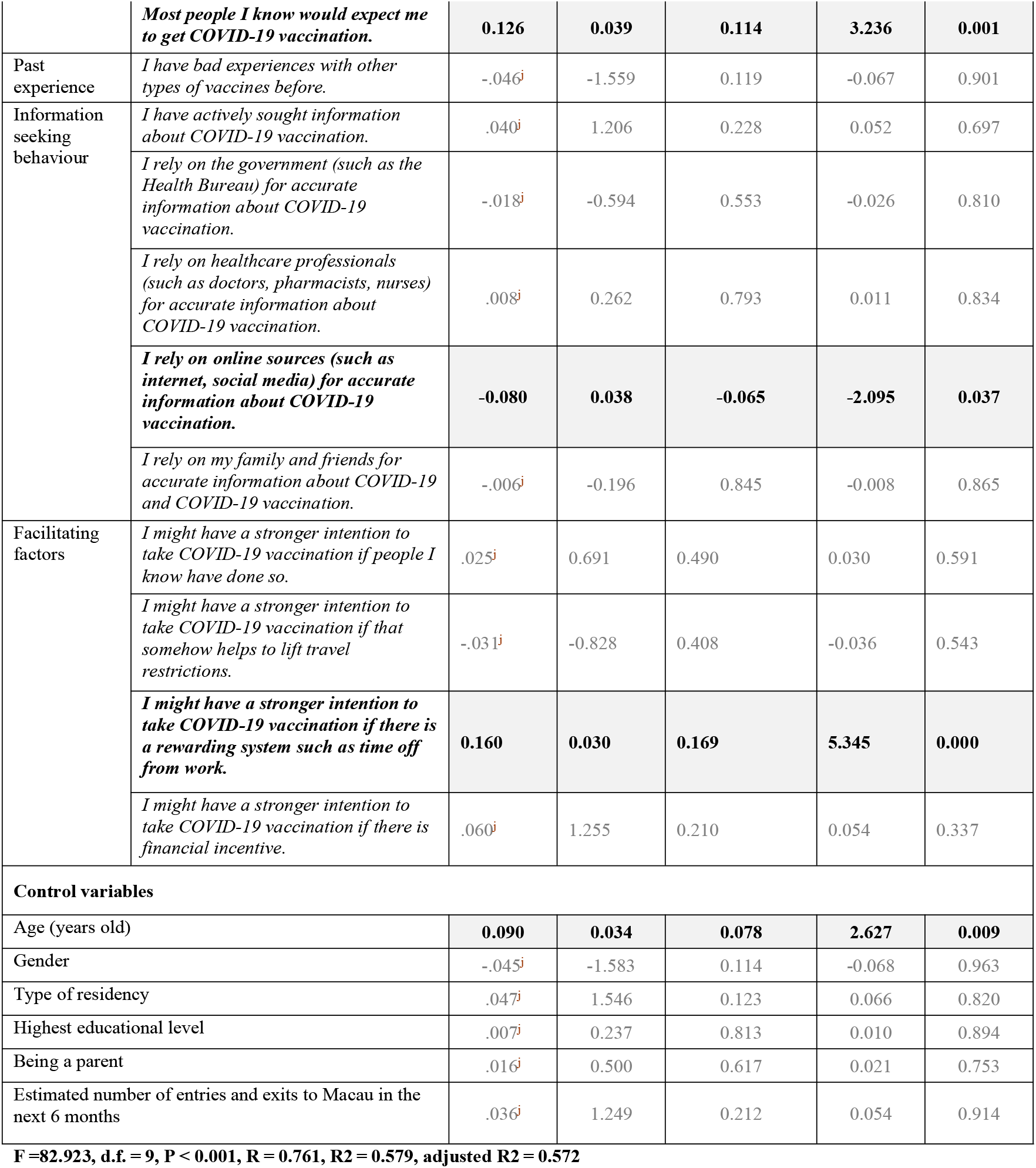
Results of multiple linear regressions.

#### Positive predictors were

(1) “*I am able to make arrangement to get COVID-19 vaccination*” in self-efficacy (β = 0.333, P <0.001); (2) “*I consider getting COVID-19 vaccination a social responsibility*” in social norm (β = 0.326, P < 0.001); (3) “*I might have a stronger intention to take COVID-19 vaccination if there is a rewarding system such as time off from work*” in cues to action (β = 0.169, P <0.001); (4) “*Most people I know would expect me to get COVID-19 vaccination*” (β = 0.114, P = 0.000) in social norm; and (5) “*The risk for me to catch COVID-19 is high*” in perceived susceptibility (β = 0.076, P <0.05).

#### Negative predictors were

(1) “*It is likely that I will have serious side-effects that I cannot bear from COVID-19 vaccination*” in response cost (β = -0.124, P < 0.001); (2) “*If I do not get a COVID-19 vaccination, I will not have to worry about the safety of the vaccine*” in maladaptive response reward (β = -0.082, P <0.05); and (3) “*I rely on online sources (such as internet, social media) for accurate information about COVID-19 vaccination*” in information seeking behavior (β = -0.065, P <0.05).

## Discussion

This is one of the few studies that investigated the intention rate of COVID-19 vaccination in areas with low incidence rate. The intention rate in Macao was 62.3%, and age, gender, type of residency, education, parental status, and having travel plans exhibited significant correlation with intention. Importantly, it was also found that the ability to make arrangement to receive the vaccine, a sense of social responsibility, an offer of time-off from work after vaccination, peer influence, and perceived susceptibility were predictors of increased intention, while concerns about vaccine safety and seeking vaccine information from online sources were predictors of decreased intention. Together with age, these predictors could explain 57.9% of the variance in vaccination intention.

The intention rate of COVID-19 vaccination in Macao (62.3%) was higher than that reported in Hong Kong (4.2% to 44.2%)^26,27^, but lower than that reported in China (88.6%-91.9%)^28,29^ and other countries (such as 94.3% in Malaysia^30^, 75.8% in Australia^31^ and 67.9% in Singapore^32^). These intention rates indicated that hesitancy and refusal about COVID-19 vaccines is a common challenge impeding the vaccine uptake across the populations.^33^ This is especially concerning for Macao considering that the low vaccination rate (around 10%) and the slow uptake since vaccination was launched in February 2021, exposing the vulnerability of the city against the spread of the virus..

### 1. Improve health literacy to counteract the negative effect of low education background

Consistent with previous findings^34,35^, respondents with lower education level indicated lower vaccination intention. On this note, the association between lower education level and poorer health was repeatedly reported^36^ and the mechanism behind the association could be explained by the concept of health literary, presenting an opportunity for intervention.^37^ Future vaccine communication strategies should consider the level of health literacy in subpopulations when considering the content and means of the promotion activities.^32^ Decreasing the complexity in communication and information processing, using infographic along with detailed information, streamlining the COVID-19 vaccination process, and involving healthcare professionals in improving public health literacy may help to mitigate the impact associated with low education level.^38,39^

### 2. Mitigate the false sense of “safety zone”

The correlation between not having travel plans and low vaccination intention was less reported in previous studies. Due to effective border control and low incidence rate in Macao, non-travelers generally felt “remote” from COVID-19 pandemic and thus perceived a low level of susceptibility despite high perceived severity affecting their intention to receive vaccination. This is worrying because for a completely naive population, a pathogen can easily spread among susceptible hosts in a rampant manner when there is sufficient exposure of susceptible hosts to infected individuals.^40^ Moreover, it would up to 2 weeks after administration of the second shot to reach the expected efficacy. Educating the public that the risks of infection would increase with the pace back to “normality” and the time lag between vaccine administration and optimal protection is crucial to help them decide about taking preventative actions in a timely manner.

### 3. Promote the social responsibility in COVID-19 vaccination and make positive action visible

Our model showed that the sense of social responsibility and peer influence could largely explain the variance in intention. According to the 2012 Global Vaccine Action Plan approved by the WHO, “*individuals and communities understand the value of vaccines and demand immunization as both their right and responsibility*”.^41^ The recognition of individual’s social responsibility should be strongly promoted within the community to leverage the power of social norm.^42^ Empowering vaccine recipients to share their personal stories and reasons for vaccination, and making positive decisions visible by, for instance, providing vaccination badges or ribbons have been recommended.^42,43^ Promoting and displaying the development momentum of social norms has been shown to be a powerful tool to shift public mentality towards health behavior change.^42,43^

### 4. Empower the public by optimizing their self-efficacy

The respondent’s ability to make arrangement to receive COVID-19 vaccine being a strong predictor reaffirmed previous findings about self-efficacy.^44,45^ To better leverage self-efficacy, the convenience of getting the vaccine could be enhanced. At present, mass vaccination sites have been employed to allow high throughput, and tailored vaccine-administration software systems enabled user-friendliness in making prior booking and onsite enrolment.^46^ Group-based, out-reach vaccination sites (e.g. corporate and universities) to reach people in their communities were also initiated. To bring vaccination services to the neighborhood, mobile vaccination trucks may be employed in future campaigns.^47^ Importantly, the “customer journey” should also be reviewed and streamlined consistently to promote vaccine recipient’s pleasant experiences. Increasing vaccination capacity to cater for increased vaccine demand, onsite registration and appointment rescheduling, as well as reminding system for the second dose and follow-up for no-shows should also be considered.^48^

### 5. Prioritize vaccination incentives (vaccination certificates and time off work, and money?)

Most people agreed that they would be more willing to receive COVID-19 vaccination should there be incentives, and preferred easing travel restrictions with vaccination and time off work over monetary incentives. Incentivizing individuals with vaccination certificates is crucial for social good, and the implications for the “recovery” of economic and socially activities are profound. Regarding time-off from work after vaccination, people might experience such side effects as fever, tiredness and muscle pain which might last for about 1-2 days.^49^ As such, as the US-CDC recommended, it is important for employers to take strategies that appropriately help their employees to manage side effects after vaccination. In contrast, the evidence about monetary incentives in changing health behavior is mixed.^50,51^ Moreover, it is difficult to determine the amount of money sufficient enough to drive people’s decision about vaccination at a population level, and yet such action would be highly resource consuming of which effectiveness remained questionable.^52^

### 6. Educate the public about scientific-based risk and benefit analysis about COVID-19 vaccines

Concerns about vaccine safety and seeking COVID-19 vaccine information from internet were found to be negative predictors of COVID-19 vaccination as reported previously.^53,54^ Misinformation regarding the vaccine safety and lack of advanced vaccination knowledge can easily contribute to anxiety, leading to overestimating possible side effects.^55,56^ On this note, there is a need for transparency and to answer concerns about the speedy development and safety of CIVD-19 vaccines.^57^ Proactive surveillance of misinformation should be reinforced. To share clear and accurate messages to otherwise unreachable population subgroups, the government may partner with trusted messengers (e.g. respected community-based groups and non-governmental organizations, opinion leaders).^32^ Less than half of the respondents considered healthcare professionals their major source of vaccine information, which indicated under-utilization of healthcare resources. Rather, healthcare professionals may be empowered to strengthen their capacity in having empathetic vaccine conversations, addressing myths, and providing vaccine information tailored to their patients.^57^

## LIMITATIONS

This study had a number of limitations. Firstly, using an online platform to invite participants and operating the survey online might have induced sampling. People with lower information technology literacy might also be under-represented in this study. By the nature of cross-sectional study, no causal inferences could be made between the key factors and the intention. There is also the timeliness to the study findings as people’s intention to receive COVID-19 vaccination is bound to fluctuate with the spread of the virus, new evidence about vaccine safety and efficacy, emergence of new variants, etc. Lastly, the vaccination intention may be affected by factors not explored in this study. Follow-up studies to identify influencing factors more comprehensively, and stratified studies to determine the differences in the impact of such factors on various subgroups would be necessary for precision vaccination strategy design.

## CONCLUSION

This study reaffirmed that low intention to receive COVID-19 vaccination is a common challenge in the combat of the pandemic. Multi-component strategies that address various factors affecting intention are needed to formulate effective interventions. Health literacy, vaccination convenience, social responsibility, reasonable incentives and well-informed risk and benefit analysis are recommended consideration for future vaccination campaigns.

## Data Availability

Data is available upon request.

## Author Contributions

COLU had full access to all the data in the study and takes responsibility for the integrity of the data and the accuracy of the data analysis. Concept and design: COLU and BY. Acquisition, analysis, or interpretation of data: All authors. Drafting of the manuscript: COLU. Critical revision of the manuscript for important intellectual content: All authors. Statistical analysis: BY. Obtained funding: COLU, YH.

## Conflict of Interest Disclosures

None reported.

## Funding/Support

The Research Fund of University of Macau (MYRG2019-00038-ICMS)

## Role of the Funder/Sponsor

The funders had no role in the design and conduct of the study;collection, management, analysis, and interpretation of the data; preparation, review, or approval of the manuscript; and decision to submit the manuscript for publication.

## Notes

### Competing Interest Statement

The authors have declared no competing interest.

### Author Declarations

The project has been approved by the Panel on Research Ethics of the University of Macau in May 2021 (SSHRE21-APP018-ICMS)

